# The Infectious Nature of Patient-Generated SARS-CoV-2 Aerosol

**DOI:** 10.1101/2020.07.13.20041632

**Authors:** Joshua L. Santarpia, Vicki L. Herrera, Danielle N. Rivera, Shanna Ratnesar-Shumate, St. Patrick Reid, Paul W. Denton, Jacob W.S. Martens, Ying Fang, Nicholas Conoan, Michael V. Callahan, James V. Lawler, David M. Brett-Major, John J. Lowe

**Affiliations:** University of Nebraska Medical Center; National Strategic Research Institute; University of Nebraska Omaha; University of Illinois at Urbana-Champaign; Harvard University

## Abstract

Severe acute respiratory syndrome coronavirus 2 (SARS-CoV-2) transmission causing coronavirus disease 2019 (COVID-19) may occur through multiple routes. We collected aerosol samples around six patients admitted into mixed acuity wards in April of 2020 to identify the risk of airborne SARS-CoV-2. Measurements were made to characterize the size distribution of aerosol particles, and size-fractionated, aerosol samples were collected to assess the presence of infectious virus in particles sizes of >4.1 µm, 1-4 µm, and <1 µm in the patient environment. Samples were analyzed by real-time reverse-transcriptase polymerase chain reaction (rRT-PCR), cell culture, western blot, and transmission electron microscopy (TEM). SARS-CoV-2 RNA was detected in all six rooms in all particle size fractions (>4.1 µm, 1-4 µm, and <1 µm). Increases in viral RNA during cell culture of the virus from recovered aerosol samples demonstrated the presence of infectious, replicating virions in three <1 µm aerosol samples (P<0.05). Viral replication of aerosol was also observed in the 1-4 µm stage but did not reach statistical significance (0.05<P<0.10). Western blot and TEM analysis of these samples also showed evidence of viral proteins and intact virions. The infectious nature of aerosol collected in this study further suggests that airborne transmission of COVID-19 is possible, and that aerosol prevention measures are necessary to effectively stem the spread of SARS-CoV-2.

## Introduction

Given the high likelihood of continued circulation of coronavirus disease 2019 (COVID-19) and the uncertain timeline for widespread vaccine-imparted immunity, optimizing infection prevention and control (IPC) practices remains a high priority for limiting transmission of Severe Acute Respiratory Syndrome coronavirus 2 (SARS-CoV-2) in healthcare settings and communities globally. Appropriate IPC protocols depend upon a robust understanding of mechanisms of transmission and relative risk of environmental and personal exposures. Data regarding SARS-CoV-2 infectivity suggests human-to-human close-proximity spread described by the classical definitions of droplet or direct contact transmission are involved in the spread of the disease^1-3^. The U.S. Centers for Disease Control and Prevention (CDC) currently recommends that healthcare workers use N-95 respirators (or better) if available when providing care for COVID-19 patients, but guidance states that the contribution of small particle aerosols to transmission remains unclear^4^, despite mounting evidence suggesting the possibility of airborne transmission^5-8^.

In order to classify an infectious disease as airborne, it must be transmitted via aerosol particles less than 5 µm in diameter. There are several lines of evidence that must be demonstrated in order to establish airborne transmission: infectious aerosol (<5 µm in diameter) must be produced by ill individuals; the infectious aerosol must be stable long enough to expose another person; and if inhaled, the viral aerosol must be capable of causing infection. Experimentally generated aerosols of SARS-CoV-2 have been shown to retain infectivity for several hours in the absence of sunlight^9-10^, and extensive contamination of COVID-19 patient care areas with SARS-CoV-2 genomic RNA suggest that aerosol dissemination of virus may occur ^11-13^. Additionally, recent clusters of COVID-19 cases linked to a church choir practice in Washington and a restaurant in Wuhan are suggestive of airborne transmission^14-15^. Finally, the primary receptor of SARS-CoV-2 for infection is understood to be ACE2^16^, which is expressed throughout the human respiratory tract, indicating that inhalation would be compatible with disease acquisition.

In an April 1, 2020, letter to the White House Office of Science and Technology Policy, the National Academy of Medicine Standing Committee on Emerging Infectious Diseases, and 21st Century Health Threats recommended further research into the role of infectious aerosols in the spread of SARS-CoV-2^17^. The work presented here demonstrates that SARS-CoV-2 RNA exists in respired aerosols; that aerosols containing SARS-CoV-2 RNA exist in particle modes that are produced during respiration, vocalization, and coughing; and that a fraction of the RNA-containing aerosols <5 µm in diameter contain infectious SARS-CoV-2 virions. The infectious nature of aerosol collected in this study, taken with the other lines of evidence presented, illustrates that airborne transmission of COVID-19 is possible, and that infectious aerosol may be produced without coughing.

## Methods

### Sample Collection

Aerosol sampling was conducted around six patients in five rooms in two wards on three separate days in April of 2020 (Table S1). An Aerodynamic Particle Sizer Spectrometer (APS 3321; TSI, Inc., Shoreview, MN) was used to measure aerosol concentrations and size distributions from 0.542 µm up to 20 µm. A NIOSH BC251 sampler^18^ was used to provide size-segregated aerosol samples for both rRT-PCR and culture analysis. The BC251 is a dry air sampler that when operated at 3.5 Lpm provides three stages of size segregated aerosol, >4.1 µm particles collected onto a 15 mL conical tube, 1-4 µm collected onto a 1.5 mL conical tube, and a 37 mm filter that collects particles <1 µm. In these studies, a gelatin filter (Sartorius, GmbH) was used in the final stage for preservation of viral integrity^19^. During these studies, APS measurements were taken at 1-minute intervals for 30 minutes in each room, with the exception of Room 5C where loss of battery power to the laptop collecting the data aborted sampling after 10 minutes. The BC251 sampler was run concurrently with the APS for 30 minutes in each room at the foot of each patient’s bed. A blank 37 mm gelatin filter was used as a control for each day to monitor for any cross-contamination of samples during handling and processing.

### Sample Recovery, RNA Extraction and Real-Time Reverse Transcriptase PCR

Recovery of the samples collected using the BC251 aerosol sampler was performed by addition of 5 mL of sterile phosphate buffered saline (PBS) to the 15 mL conical tube, 1 mL of sterile PBS to the 1.5 mL conical tube, and by placing the 37 mm gelatin filter from the last stage in a 50 mL conical tube and dissolving into 10 mL of sterile PBS.

RNA extractions were performed on 400 uL of initial sample using a Qiagen DSP Virus Spin Kit (QIAGEN GMbH, Hilden, Germany). A negative extraction control (no sample added) was included with each set of extractions. Samples were eluted in 50uL of Qiagen AVE Buffer. rRT-PCR was performed using Invitrogen Superscript III Platinum One-Step Quantitative rRT-PCR System. Each rRT-PCR run included a positive synthetic DNA control and a negative, no template, control of nuclease free water. Reactions were set up and run with initial conditions of 10 minutes at 55°C and 4 minutes at 94°C then 45 cycles of 94°C for 15 sec and 58°C for 30 seconds, on a QuantStudio™ 3 (Applied Biosytems™, Inc) with the reaction mix and primers and probe targeting the E gene of SARS-CoV-2 ^13,20^ (details provided in Table S2).

To quantify the virus present in each sample from the measured Ct values obtained from the rRT-PCR, a standard curve was developed using RNA extracted from a known quantity of SARS-CoV-2 virus (BEI_ USA-WA1/2020) cultivated in Vero-E6 cells (using the same method described below for environmental samples). A 6-log standard curve was run in duplicate beginning at a concentration of 1×10^2^ TCID_50_/mL, as determined by media tissue culture infectious dose (TCID_50_) assay. The data were fit with the exponential function:

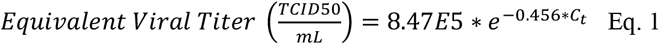

In the equation, *C*_*t*_ is the cycle time where amplification is definitively above the background. The same cycle threshold was used for all rRT-PCR runs in this study. Equation 1 was used to convert measured *C*_*t*_ values to an equivalent concentration of virus. With this conversion, Amplification at 45 cycles indicates equated 5 TCID_50_ in the reaction volume. The resulting limit of detection for the rRT-PCR was ∼20 TCID_50_/mL of extracted sample. The average and standard deviation concentrations were calculated in triplicate for each sample.

### Cell Culture and Detection of Viral Replication

Vero E6 cells were used to culture SARS-CoV-2 virus from environmental samples. The cells were cultured in Dulbeccos’s minimal essential medium (DMEM) supplemented with heat inactivated fetal bovine serum (10%), Penicillin/Streptomycin (10,000 IU/mL &10,000 µg/mL) and Amphotericin B (25 µg/mL). For propagation of infectious virus from the aerosol samples, 100 µL of undiluted samples were added to 24-well plates containing confluent monolayers of Vero E6 cells and grown at 37 ° C with 5% CO_2_. To look for evidence of viral replication, RNA was extracted from 140 uL of supernatant from days 1 and either day 5 or 6 of incubation, using the QIAamp Viral RNA Mini Kit (QIAGEN GMbH, Hilden, Germany) and rRT-PCR was performed on these samples following the methods described for environmental samples. The percent increase from day 1 to day 5 or 6 was calculated for all samples to look for evidence of viral replication in each sample. Definitive replication was considered to occur for rRT-PCR samples in which a significant increase in RNA was detected in the supernatant (student’s t-test, Microsoft Excel, P<0.05). After six days, all cell supernatants and lysates were harvested. For samples with statistically significant evidence of replication, subsequent examination of the samples was performed by electron microscopy and western blot assay. Western blots were generated by harvesting cell lysates in RIPA lysis buffer. Twenty uL of lysate was combined with a 2X reducing sample buffer (Invitrogen), boiled and subjected to sodium dodecyl sulfate polyacrylamide gel electrophoresis (SDS-PAGE) analysis followed by Western blot using the antibody against SARS-CoV N protein and glyceraldehyde 3-phosphate dehydrogenase (GAPDH) (Abcam). For electron microscopy, samples were fixed, processed, sectioned, and inspected (see Table S3) on a Tecnai G2 Spirit TWIN (Thermo Fisher Scientific) at 80kV.

### Aerodynamic Data Analysis

APS measurements were taken at one-minute intervals and exported as mass concentration in *dM/dlogD*_*p*_ per size bin, assuming particles of unit density. The first bin of the APS data files (<0.523 μm) was discarded during data analysis. An average concentration across all bins for all one-minute measurements was calculated. The averaged raw distributions were then parameterized into two log-normally distributed aerosol modes. The parameterized distributions were assumed to be lognormal and the Solver function in Microsoft Excel was used to calculate the fit parameters, *M*, the total mass concentration, *d*_*pg*_, the mass median aerodynamic diameter, and *σ*_*g*_ the geometric standard deviation for each mode by using a minimization of the sum of least square errors between the raw distribution and parameterized distributions^21^. The total mass of each mode contributing to the raw distributions was summed from 0.542 to 0.97 μm, to 3.79 μm, and 4.07 to 9.85 μm to reflect the size ranges overlapping with the different stages of BC251 aerosol sampler. The total concentration in each size bin was normalized such that both the APS parameterized modes and modes obtained from the rRT-PCR analysis performed off the BC251 samples could be compared directly.

## Results

In all six rooms, rRT-PCR indicated the presence of viral RNA in all three stages of the BC251 sampler (Figure 1; Table S4). Furthermore, exposure of recovered samples to Vero-E6 cells demonstrated statistically significant viral growth (95% confidence based on P<0.05) after six days (five days in the case of the submicron sample from room 5A) in 3 of the 18 samples (7B, 5A and 5C; Figure 1). Two of the 1-4 µm samples demonstrated viral growth, between 90% and 95% confidence (7B and 5C; Figure 1). Cells from these five samples were subsequently examined via western blot for the presence of viral protein production and transmission electron microscopy for the presence of intact SARS-CoV-2 virions. The presence of SARS-CoV-2 was observed via western blot for all but one of the samples (<1 μm, Room 7B) with statistically significant evidence of replication, by rRT-PCR (Figure 2). Intact virus was observed via TEM in the submicron sample from Room 5C (Figure 3), further indicating active viral replication in that sample.

**Figure 1.**
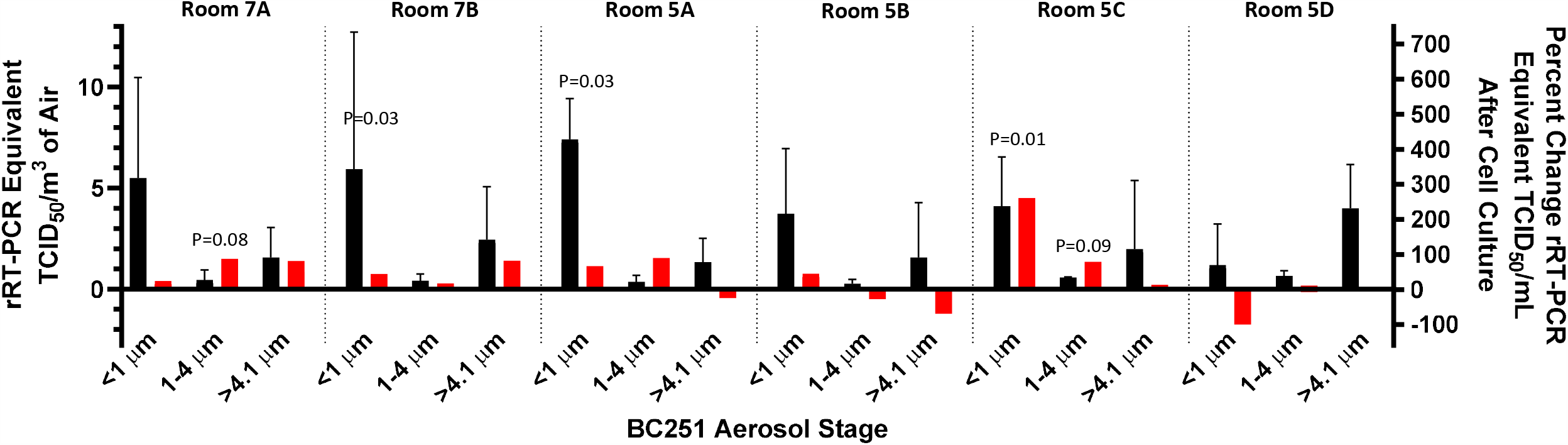
Measured Airborne RNA Concentrations and Associated Percent Change in Viral Copies in Cell Culture. Measured RT-PCR equivalent TCID50/cm^3^ (black) indicate in initial viral RNA concentrations while the percent change viral copies (red) indicates viral replication in cell culture (or lack thereof). Error bars indicate the standard deviation of the RT-PCR-derived concentrations (for air concentration) and calculated measurement uncertainty (for percent change in cell culture). The P-value comparing the initial and final PFU/mL measurements is shown if the change is positive and the P-value is less than 0.1. Three of the six submicron filter samples indicated an increase in cell culture that was significant (p<0.05) based on the Students T-Test (7B, 5A and 5C. Two of the 1-4 um samples had P-values less than 0.1, but not less than 0.05 (7A and 5C). None of >4.1 um samples demonstrated statistically significant replication.

**Figure 2.**
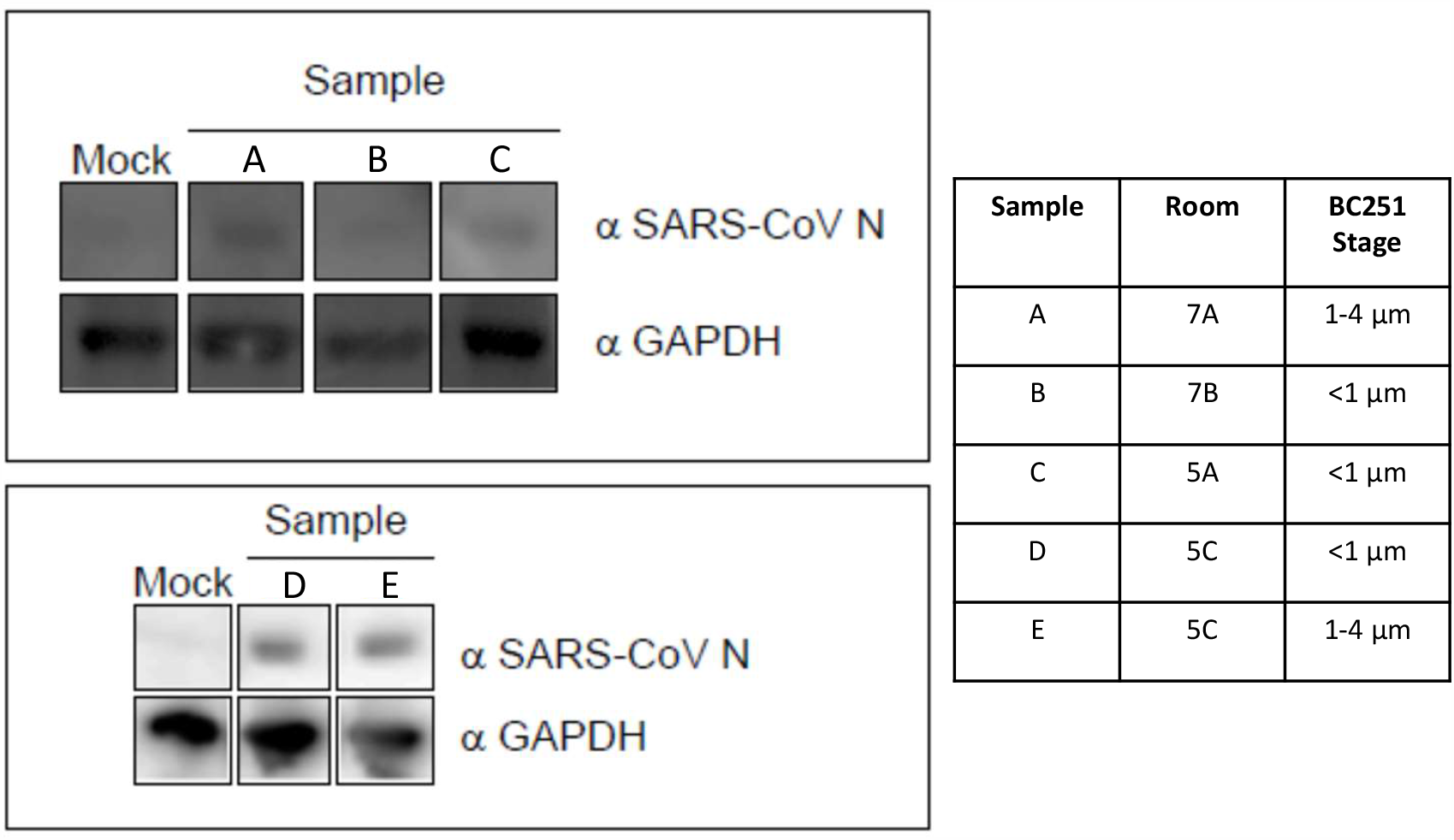
Protein expression of SARS-CoV-2. Cell protein lysates were prepared from the indicated cultured samples and subsequently probed by western blot with a mouse monoclonal anti-SARS nucleocapsid protein (SARS-CoV N) antibody and an anti-GAPDH loading control antibody.

**Figure 3.**
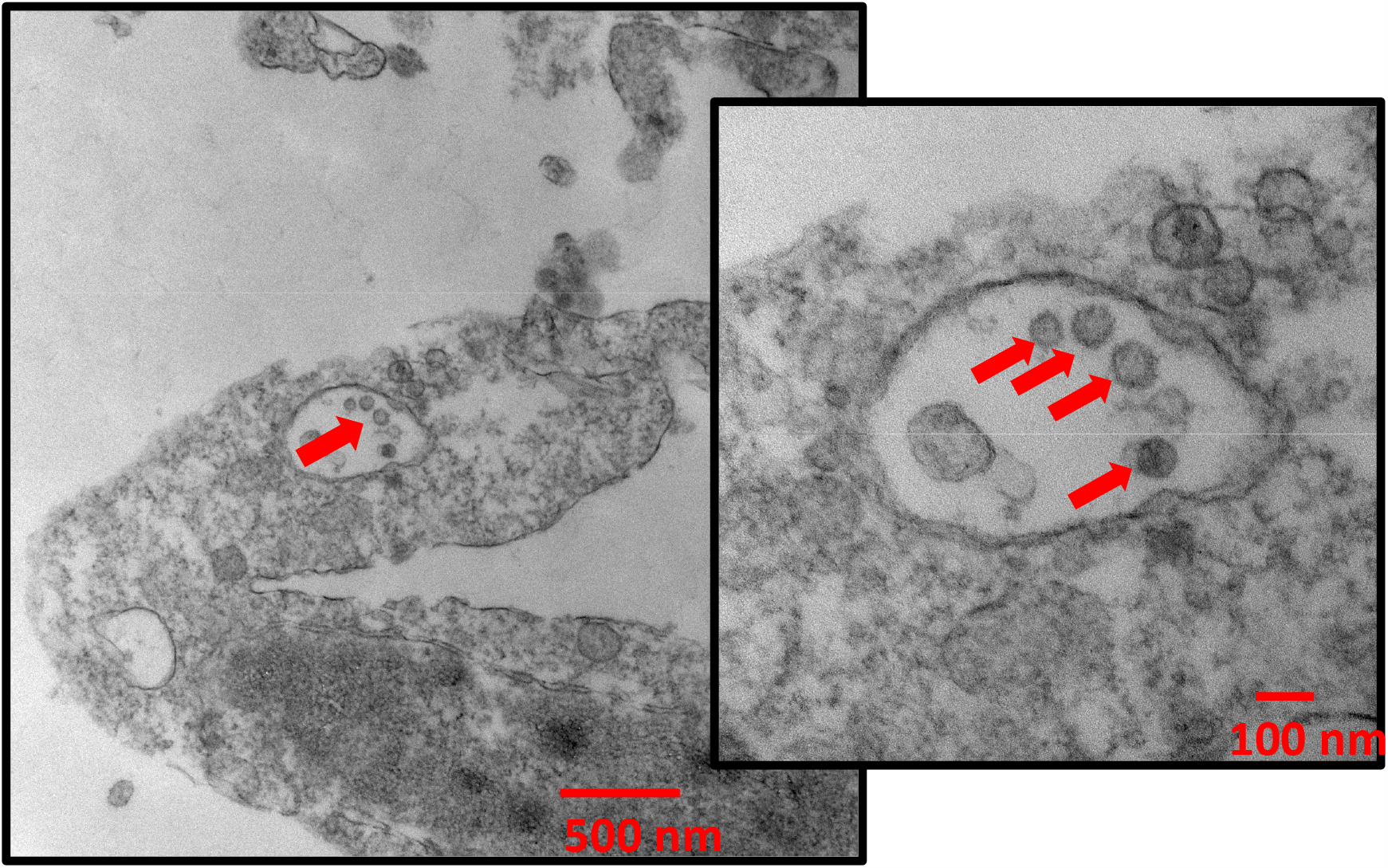
Electron micrographs of SARS-CoV-2 virions cultivated from the sub-micron filter from Room 5C.

Two log-normal modes were calculated from the measured APS aerosol mass distributions (Figure 4 and Table S5). The rooms from ward 5 all have very similar distributions and parameterized modes, while the rooms in 7 have much lower concentrations and broader size distributions than those in ward 5 (Figure 4). The mean diameter of the small mode distributions is between 0.64 µm and 0.80 µm for all the rooms sampled (Table S5), while the width (GSD) of the distribution varied between 1.17 and 1.30. The large mode distributions had more variability both in terms of mean diameter and width (Table S5).

**Figure 4.**
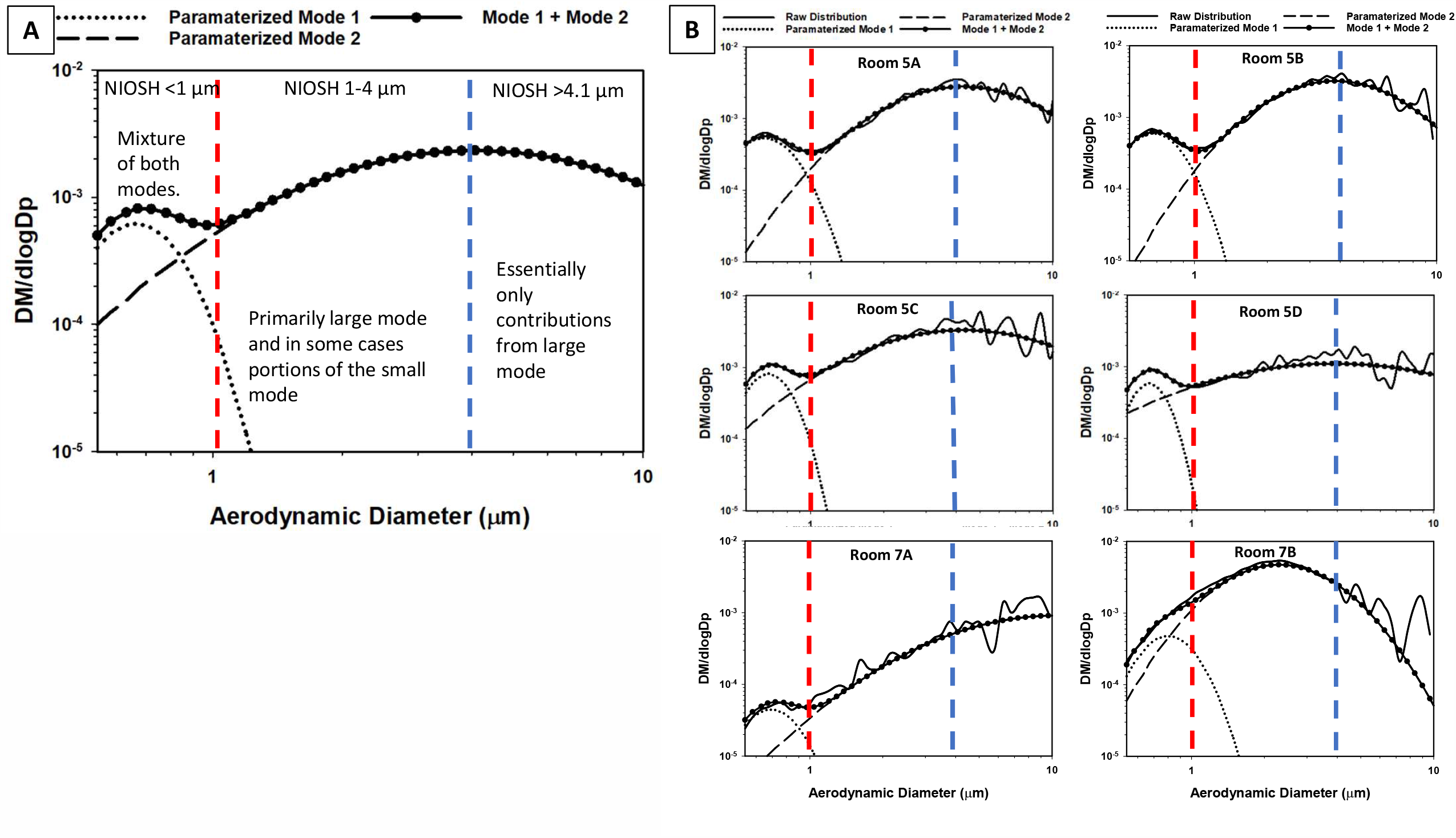
Aerosol Mass Distributions. (assuming unit density particles), including measured (solid lines), the parameterized small mode (dots with no line), parameterized large mode (dashed line) and the combination of the two modes (heavy dots with line) for the average of all rooms in ward 5 (A), and each individual room in the study (B). The red and blue lines indicate the cut-off size for the BC251 sampler, and indicate which fraction of each of the total aerosol and each mode was collected by the BC251 sampler.

## Discussion

This study sought to characterize the presence of SARS-CoV-2 in particles consistent with the potential to result in aerosol transmission between patients. Although not all particles measured by the APS may be attributable to patient extrusions, increases in particle count while measurements were being taken using the APS were anecdotally observed to occur when patients were talking and coughing. The aerosol modes observed in this study were compared to those from previous observations of human aerosol production during respiratory activities. The small aerosol mode, with a mean diameter between 0.64 and 0.80, is consistent with particles found in exhaled breath in previous studies^22-24^. This mode of aerosol was observed in all manner of human respiration including breathing, vocalization and coughing^22^ and has been attributed to particles produced deep in the bronchial region, referred to as the Bronchiolar Fluid Film Burst (BFFB) mechanisms^23^. Particles in the larger modes observed in this study are more consistent with those produced in the larynx during vocalization and coughing^25^.

Observation of fine mode aerosol particles containing infectious SARS-CoV-2 particles leads to several general observations about the potential transmission of SARS-CoV-2. The results of this study, along with the evidence of the stability of SARS-CoV-2 in aerosol^9-10^ and that SARS-CoV-2 infects respiratory tissue^16^ provide indications that SARS-CoV-2 may be transmitted via the airborne route. Furthermore, fine mode aerosols are generally produced in the bronchi and the larynx across a wide range of respiratory activities, which is in agreement with previous studies in which SARS-CoV-2 aerosol are observed in the absence of coughing and other symptoms^13^ as well as in reports of disease transmission by asymptomatic individuals^14-15^.

The infectious SARS-CoV-2 submicron aerosols detected in this study were collected in the last stage of the BC251 sampler. This sampler was not originally intended to collect and preserve the integrity of SARS-CoV-2 in aerosol, and there have not yet been studies to determine how effective this approach might be at preserving the infectivity of the virus. In the first two stages of the sampler, the virus is sampled into two dry conical tubes using inertial impaction. It is possible that this collection mechanism may be impacting the integrity of the virus collected in the 1-4 µm and the >4.1µm stages. For this study, we employed gelatin filter in the final stage of the BC251 to help preserve intact virus during collection and notably it was in this stage that viral replication was observed. Therefore, it can be said that infectious SARS-CoV-2 exists in particles <1 µm and may exist in particles up to 4 µm, but it cannot be said that they do not exist in particles larger than 4 microns, due to the limitations of the collection technique.

A comparison of the APS and BC251 aerosol data (Table S6) is suggestive of a relationship between the measured mass distributions and viral RNA concentrations from the different stages of the BC251 sampler. However, the small number of data points (6, in all cases), does not allow the determination of a statistically relevant relationship. Despite that, examination of the observed APS modes against the size bins of the BC251 sampler and comparison to studies of human respiratory emissions does offer valuable insight into the source of RNA-containing and infectious aerosol present in these samples. SARS-CoV-2-containing aerosols collected in the submicron filter of the BC251 is likely attributable to a combination of the small and large modes that were fit to the APS data, with the dominant fraction of the aerosol mass attributable to the small mode. This would indicate that the portion of aerosol containing both RNA and infectious aerosol that were identified in three of the rooms likely had their origins in the bronchial region of those patients. The 1-4 µm BC251 stage was dominated by the large mode, in which the human associated fraction is most consistent with particles produced in the larynx, and are produced more heavily during vocalization and coughing. The largest size bin of the BC251 (>4.1 µm) would have been associated almost exclusively with the large aerosol mode observed in the APS, and possibly other particles and droplets not measured by the APS, such as those produced in the oral cavity that are generally much larger^25^. Although viral RNA was consistently observed in aerosol in this largest stage, no infectious aerosol were recovered from it.

## Conclusion

Our results demonstrate that SARS-CoV-2 RNA exists in respired aerosols less than 5 µm in diameter; that aerosols containing SARS-CoV-2 RNA exist in particle modes that are produced during respiration, vocalization, and coughing; and that some fraction of the RNA-containing aerosols contain infectious virions (Table S7).

This study supports the use of efficient respiratory protection and airborne isolation precautions to protect from exposure to fine SARS-CoV-2 aerosol when interacting with infected individuals, regardless of symptoms or medical procedure being performed.

Given the prospect of continued widespread circulation of COVID-19, and recent work highlighting the relative importance of airborne transmission of COVID-19^5^, it is crucial that evidenced-based IPC practices are promoted and implemented to limit the transmission of SARS-CoV-2 in healthcare, community and industry settings. Given the infectious nature of aerosol collected in this study, taken with the other lines of evidence presented, further suggests that airborne transmission of COVID-19 is possible, and that aerosol prevention measures should be implemented to effectively stem the spread of SARS-CoV-2, particularly in crowded settings.

## Data Availability

All data is available in the main text or the supplementary materials

## Acknowledgements

The authors would like to thank the Electron Microscopy Core Facility (EMCF) at the University of Nebraska Medical Center for technical assistance. The EMCF is supported by state funds from the Nebraska Research Initiative (NRI) and the University of Nebraska Foundation, and institutionally by the Office of the Vice Chancellor for Research. We also thank William Lindsley for the loan of the NIOSH BC251 samplers. Finally, we would also like to thank the nurses and doctors who helped facilitate access to the patient rooms for sampling, and to the patients themselves for allowing us access to their rooms during their stay at Nebraska Medicine.

## Ethical Review

This study was conducted with permission from the University of Nebraska Medical Center and Nebraska Medicine as a part of a quality assurance/quality improvement study on isolation care. This activity was reviewed by Office of Regulatory Affairs at the University of Nebraska Medical Center and it was determined that this project does not constitute human subject research as defined by 45CFR46.102.

## Data and materials availability

All data is available in the main text or the supplementary materials

## Author Contribution Statement

The study was conceived by JLS. Samples were collected by JLS and JJL. Samples were processed by DNR and VLH. rRT-PCR of all samples was performed by VLH. Cell culture work was performed by SPR with assistance by PWD and JWSM. YF generated the antibodies. NC performed EM. JLS and SRS performed aerosol analyses. DMB aided with patient interactions. JLS, JJL, SPR, MVC and JVL wrote the manuscript with contributions from all authors.

